# Prevalence of subjective cognitive decline among older Multiracial adults, 2019-2023

**DOI:** 10.1101/2025.06.23.25329943

**Authors:** Tracy Lam-Hine, Michelle C. Odden, Bryan D. James, David H. Rehkopf

## Abstract

**Background:** The Multiracial (two or more races) population is the fastest-growing U.S. racial and ethnic group but remains underrepresented in cognitive aging research. No national estimates exist for subjective cognitive decline – a self-reported indicator of worsening memory that is associated with increased dementia risk – among Multiracial older adults. We used descriptive epidemiologic methods to estimate prevalence in this group.

**Methods:** We pooled Behavioral Risk Factor Surveillance System (BRFSS) data from 2019–2023 from U.S. states and territories that administered the optional cognitive decline module. Our analytic sample included 546,371 adults aged 50 years or older. We estimated crude and age/sex-adjusted prevalence of subjective cognitive decline by self-identified race and Hispanic ethnicity using survey-weighted logistic regression with predictive marginal standardization. We stratified adjusted prevalence estimates by state.

**Results:** Adjusted subjective cognitive decline prevalence was highest among American Indian or Alaska Native (18.2%) and Multiracial (17.9%) adults, nearly double the prevalence observed among Asian adults (9.1%). Among Multiracial adults, state-level adjusted subjective cognitive decline prevalence ranged from 15.2% in Puerto Rico to 20.5% in New Mexico, with low geographic variation (interquartile range: 18.8%–19.6%) and no consistent regional pattern.

**Conclusions:** This study provides the first national estimates of subjective cognitive decline among older Multiracial adults, revealing a high and relatively uniform burden across states. These findings underscore the need to better characterize cognitive aging risks in this growing population and support the inclusion of Multiracial individuals in longitudinal and clinical dementia studies.

## Introduction

The Multiracial (two or more race) population is the fourth-largest and fastest-growing U.S. racial and ethnic group, with over 33.8 million adults (10.2%) selecting multiple races in the 2020 Census.^1^ Evidence suggests that Multiracial individuals share unique and diverse social experiences that may influence health and aging,^2,3^ however, Multiracial individuals remain underrepresented in cognitive aging and dementia research. Prior studies using clinical assessments and administrative data have identified racial and ethnic disparities in the prevalence of dementia and cognitive impairment; however, estimates for the Multiracial population remain unknown.^4–8^ These studies largely exclude Multiracial individuals, recategorize them into single race groups, or rely on datasets with limited race data or racial diversity. For example, Matthews and coauthors used Medicare claims data to estimate the number of current dementia cases stratified by race and ethnicity.^4^ This is the only existing study we know of that attempts to estimate prevalence among the Multiracial population (43,000 cases in 2014). However, the authors arrived at this estimate by applying the “age-sex-specific prevalence for *all* races to the total Census population of two or more races”, as the Centers for Medicare and Medicaid Services does not report these data using a Multiracial (or equivalent) category. As a result of these dataset and analytic limitations, there is little empirical understanding of the distribution of cognitive health or early indicators of dementia risk in this population.

Findings from several studies using national health statistics reveal high prevalence of several major risk factors for cognitive impairment. Omura and coauthors used 2019 Behavioral Risk Factor Surveillance System (BRFSS) data from 31 states and the District of Columbia to compare racial and ethnic differences in the prevalence of eight common modifiable risk factors for Alzheimer’s disease and related dementias, finding that Multiracial adults had the highest prevalence of depression and second-highest prevalence of current cigarette usage, binge drinking, and hearing loss among all groups studied.^9^ And using 2022 BRFSS data from 39 states, Town and coauthors found that Multiracial adults had the highest prevalence of life dissatisfaction and social isolation or loneliness and second-highest prevalence of mental stress.^10^ Given growing evidence linking stress, isolation, and loneliness to increased risk of cognitive aging and Alzheimer’s disease and related dementias,^11,12^ these studies’ results point to the importance of systematically surveilling and examining cognitive health patterns in the Multiracial population.

In contrast to clinical or administrative datasets, population-based surveys offer greater racial diversity and the potential to include underrepresented groups such as Multiracial adults in studies of cognitive decline and dementia risk. One cognitive measure commonly assessed in these surveys is subjective cognitive decline, a self-reported measure of worsening confusion or memory loss. Although subjective cognitive decline is not a clinical diagnosis and has limited sensitivity for detecting objective cognitive impairment, it is increasingly understood as a putative risk factor and is useful for making comparisons across population subgroups in public health surveillance.^13–15^ Critically, it is one of the only cognitive health indicators available in large-scale surveys that include Multiracial participants, making it a necessary starting point for examining cognitive aging patterns in this population. Prior descriptive studies of population-based datasets suggest that racial disparities in subjective cognitive decline largely mirror those in cognitive impairment, with Black, Hispanic, and Indigenous groups reporting higher prevalence than the White or Asian populations.^15–17^ Yet despite the availability of data on Multiracial people in these datasets, authors of these studies excluded them from analysis or reassigned them into an uninformative “other” racial category.

To address this gap, we used a large, nationally representative survey to estimate and compare national subjective cognitive decline prevalence among adults aged 50 and older across racial and ethnic groups, with a focus on the Multiracial population.

## Methods

The Behavioral Risk Factor Surveillance System (BRFSS) is a phone-based, cross-sectional survey assessing health and related factors among adults ages 18 and older, residing in U.S. states, participating territories, and the District of Columbia. The survey includes self-reported information on demographics, health behaviors, preventive care, and chronic conditions. Beyond the core annual survey, participating jurisdictions can elect to administer optional modules in a given year, including a cognitive decline module.^18^ For each year from 2019 to 2023, we identified all states and territories that administered the cognitive decline module, and pooled their responses to maximize sample size and representation. In the cognitive decline module, respondents answered yes or no to the following question: “During the past 12 months, have you experienced confusion or memory loss that is happening more often or getting worse?” We included all respondents aged ≥ 50 years in our analysis, and categorized respondents that answered “yes” as experiencing subjective cognitive decline.

We estimated crude and adjusted prevalence of subjective cognitive decline, incorporating survey weights to account for BRFSS’s complex survey design. We stratified crude estimates by binary sex (male/female) and 5-year age categories (50-54 to 80+ years). To control for differing age and sex distributions across racial and ethnic groups, we adjusted for binary sex and age category using predictive marginal standardization from a survey-weighted logistic regression model. We then estimated adjusted prevalence by self-identified Hispanic ethnicity (of any race) and non-Hispanic race, which BRFSS reports in seven mutually exclusive groups (American Indian or Alaska Native, Asian, Black, Native Hawaiian or Pacific Islander, Multiracial, White, another race), as well as by state of residence. Consistent with established methods guidance for descriptive epidemiologic analyses, we did not adjust for potential mediators or risk factors (e.g., depression, comorbidities), which could distort or obscure real disparities.^19^ Taylor linearization calculated standard errors for adjusted prevalence estimates. We handled missing data using multiple imputation via chained equations (20 imputations, 20 iterations) under the assumption of missing at random, and compared imputed and complete case data to assess consistency.

Following prior guidance,^20^ we also included additional demographic and health predictors in the imputation model that were not part of our main analysis (further details available in **Supplement**). We conducted all analyses in R version 4.4.0 (R Core Team, Vienna, Austria, 2024), and used ChatGPT-4o (OpenAI, 2025) to support coding review and manuscript editing. Fully reproducible code for this analysis is available at https://github.com/lamhine/brfss_scd.

The Stanford University institutional review board exempted this study as data are deidentified and publicly available.

## Results

Among 546,371 adults (46.8% male and 53.2% female) ages 50 and over, the largest racial and ethnic groups were White (70.0%), Hispanic (12.0%), Black (12.0%), and Asian (3.5%) (**Supplement**). Smaller proportions identified as Multiracial (1.5%), American Indian or Alaska Native (1.2%), Native Hawaiian or Pacific Islander (0.2%), or another race (0.8%).

Crude prevalence of subjective cognitive decline by age group followed slightly different patterns for strata of sex and racial or ethnic group (**Figure 1**). Among males, prevalence was relatively stable for individuals ages 50 to 74 (11.3%–12.7%), followed by a sharp increase at ages 75-79 (14.5%) and 80+ (17.3%). For females, prevalence was also highest in the oldest (80+, 16.1%) age groups, however, there was a modest U-shaped pattern in the intermediate age categories (60-64, 13.5%; 65-69, 12.4%; 70-74; 12.7%) not seen among males. In most age-sex-race strata, prevalence increased with age, though the shape and steepness of age gradients varied. Some groups, such as Asian males and Hispanic females, showed gradual and consistent increases across age groups. Others, including Black females and White males, displayed relatively flat patterns with only modest age-related changes. Several smaller subgroups—such as AIAN males and Multiracial females—showed more fluctuation across age groups, potentially reflecting instability due to limited sample size.

**Figure 1.**
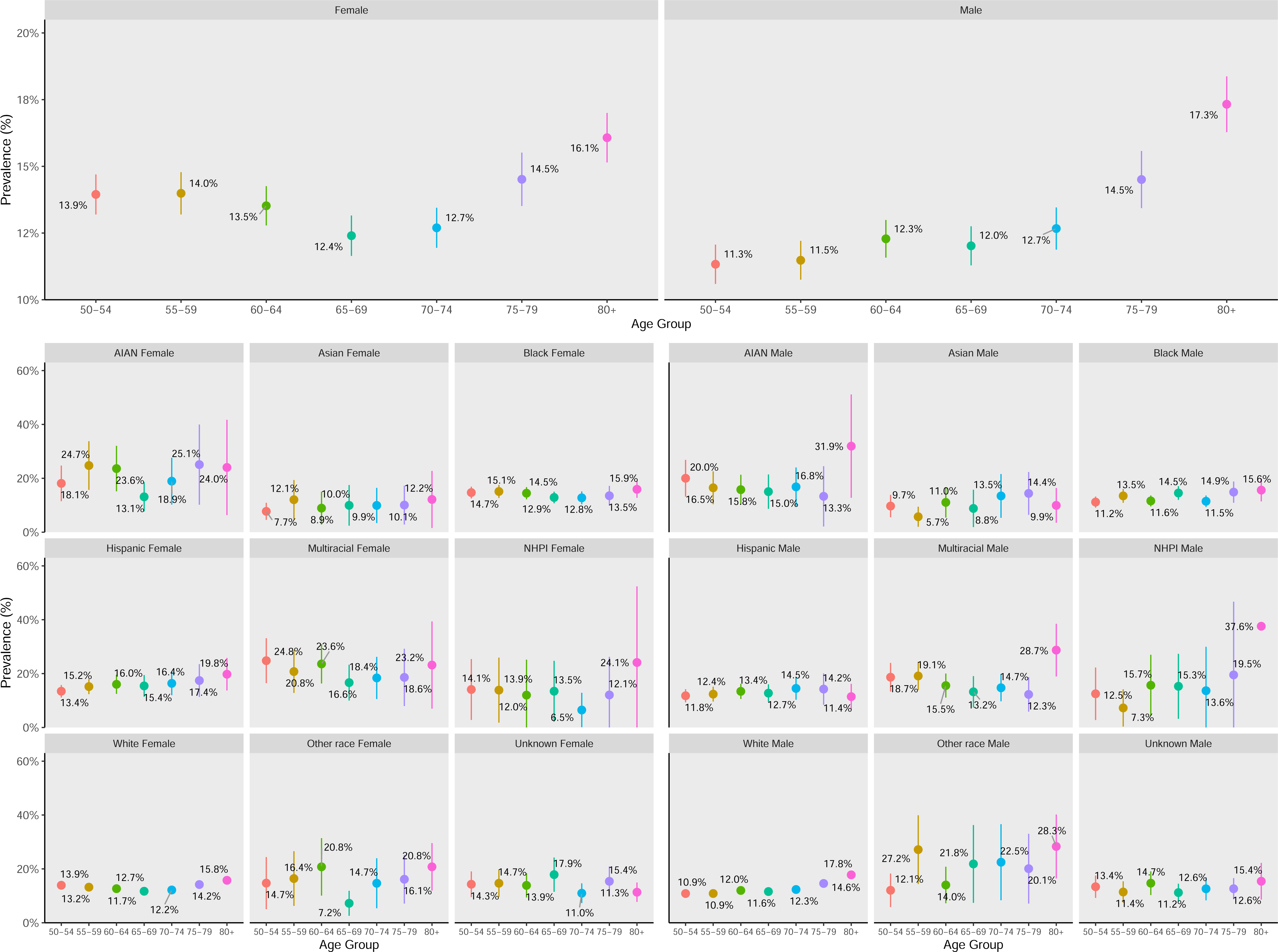
Crude prevalence of subjective cognitive decline by sex, age, and racial and ethnic groups, BRFSS 2019-2023 Abbreviations: BRFSS = Behavioral Risk Factor Surveillance System

Age- and sex-adjusted prevalence of subjective cognitive decline increased modestly for most racial and ethnic groups from 2019 to 2023, with a notable convergence in 2021 for all groups other than AIAN adults (**Figure 2**). Multiracial and AIAN adults consistently had the highest prevalence across the 5-year period, while Asian adults remained the lowest. Across the five-year period, overall adjusted prevalence was 14.1% (95% CI: 13.3%–14.9%), and was highest for American Indian or Alaska Native (18.2%, 95% CI: 15.8%–20.6%) and Multiracial (17.9%, 95% CI: 15.8%–20.0%) adults. Other groups ranged from 13.4% (Hispanic, 95% CI: 12.4%–14.4%) to 9.1% (Asian, 95% CI: 7.5%–10.6%). Multiply imputed and complete case results were similar.

**Figure 2.**
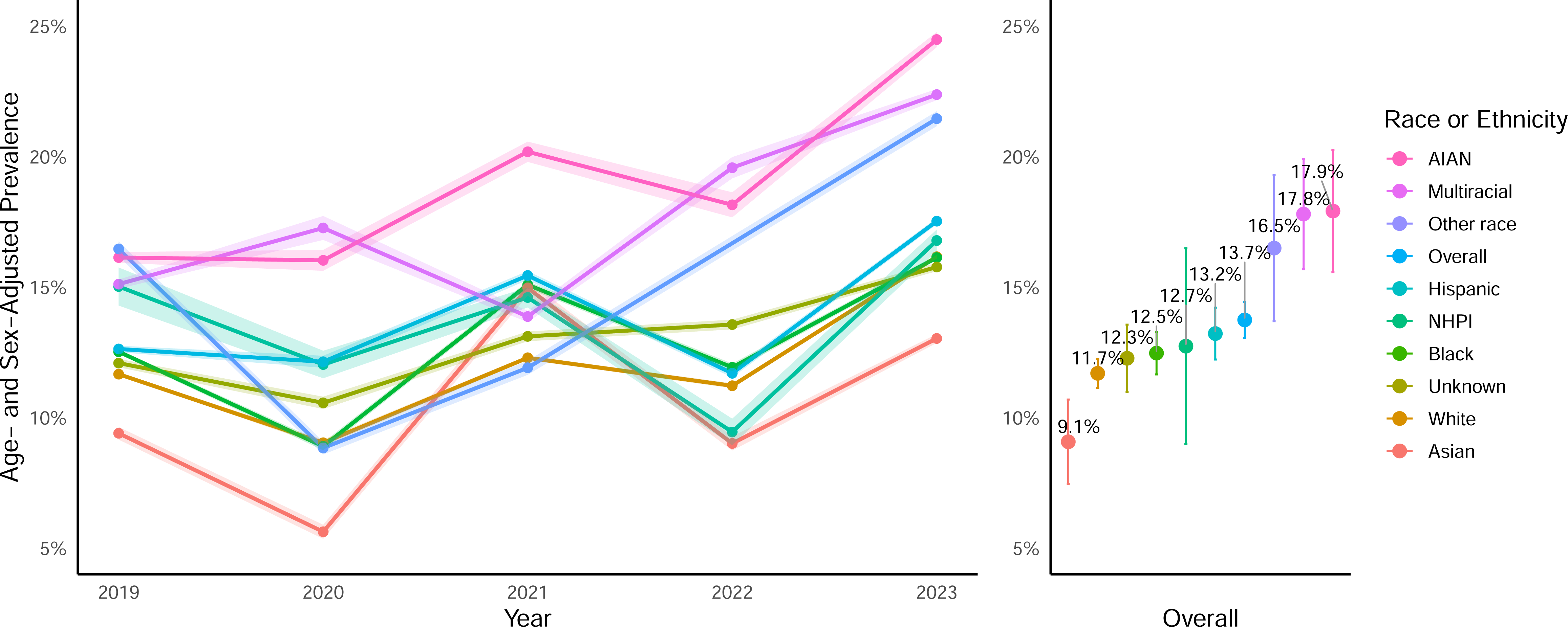
Annual and overall adjusted^a^ prevalence^b^ of subjective cognitive decline by self-identified race and ethnicity, BRFSS 2019-2023 Abbreviations: BRFSS = Behavioral Risk Factor Surveillance System; AIAN = American Indian or Alaska Native; NHPI = Native Hawaiian or Pacific Islander ^a^ Adjusted for sex and 5-year age categories using predictive marginal standardization ^b^ Error bars represent 95% CIs obtained via Taylor linearization

Adjusted subjective cognitive decline prevalence among Multiracial participants ranged from 15.2% in Puerto Rico to 20.5% in New Mexico, with low variation across states (interquartile range: 18.8%–19.6%) (**Figure 3**). No consistent geographic or regional pattern emerged, with both the highest (New Mexico: 20.5%, 95% CI: 18.6%–22.4%; Utah: 20.4%, 95% CI: 19.6%–21.1%; Louisiana: 20.2%, 95% CI: 19.2%–21.2%; District of Columbia: 20.1%, 95% CI: 19.2–21.0%; New Hampshire: 20.1%, 95% CI: 19.0–21.2%) and lowest (Puerto Rico: 15.2%, 95% CI: 13.7%–16.6%; Massachusetts: 17.3%, 95% CI: 15.8–18.7%; Illinois: 18.2%, 95% CI: 16.8%–19.6%; South Dakota: 18.4%, 95% CI: 17.7%–19.2%) prevalence estimates distributed across multiple U.S. regions.

**Figure 3.**
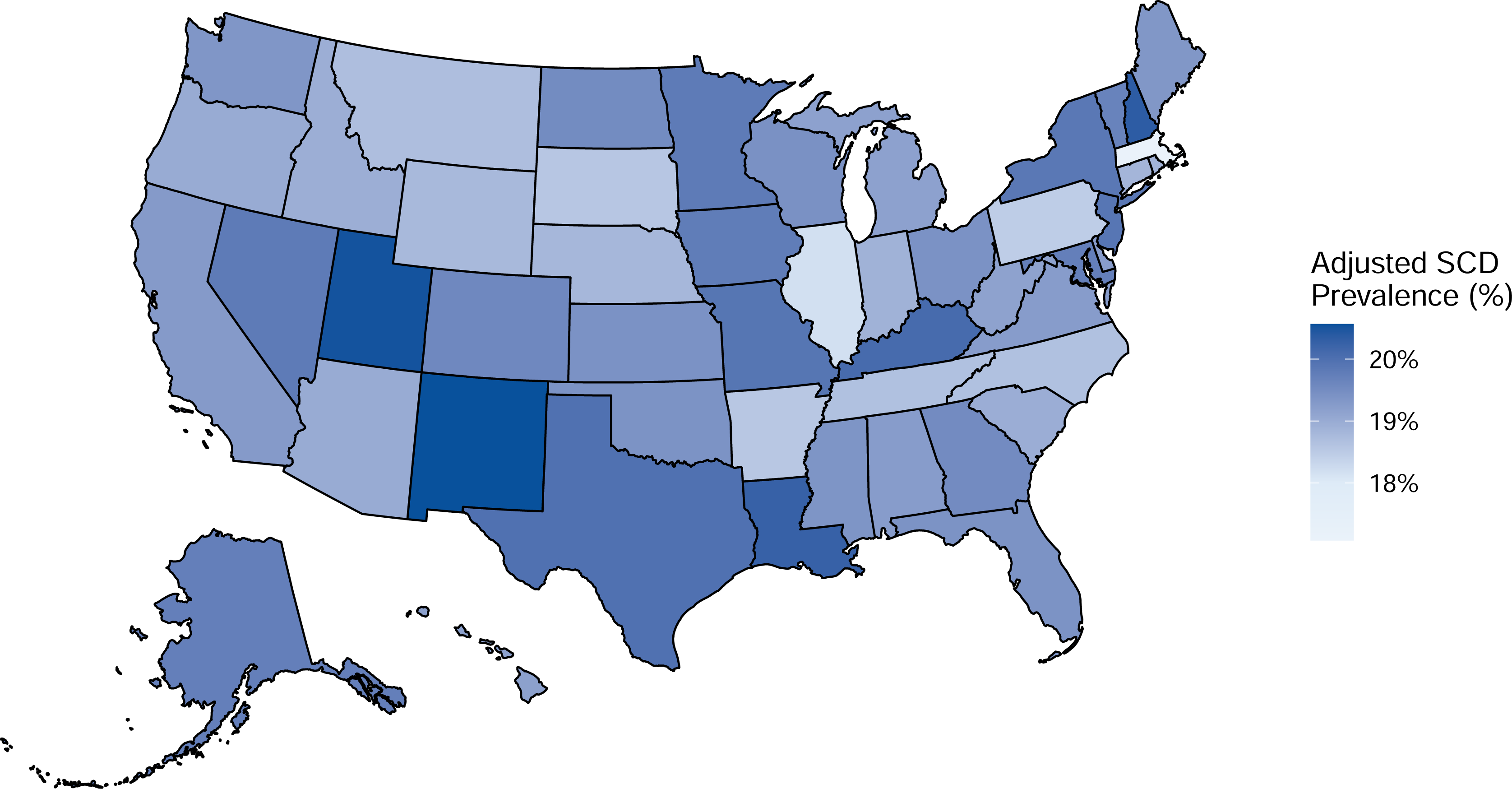
Adjusted^a^ prevalence of subjective cognitive decline among Multiracial adults ages 50+ by U.S. state^b^, BRFSS 2019-2023 Abbreviations: BRFSS = Behavioral Risk Factor Surveillance System ^a^ Adjusted for sex and 5-year age categories using predictive marginal standardization ^b^ Map displays data for the 50 U.S. States and District of Columbia; Puerto Rico (adjusted prevalence = 15.1%) not shown

## Discussion

This study provides the first national estimates of subjective cognitive decline prevalence among older Multiracial adults, revealing a high burden comparable to American Indian or Alaska Native adults and nearly twice that of Asian adults. In contrast to prior national analyses reporting substantial geographic variation and lower overall subjective cognitive decline prevalence,^21^ we found consistently high subjective cognitive decline prevalence among Multiracial adults across states with relatively modest variation. Although subjective cognitive decline is not a clinical diagnosis and has limited sensitivity for detecting objective cognitive impairment, it is one of the only cognitive health indicators available in large, racially diverse surveillance systems such as BRFSS.^13–15^ Its utility for subgroup comparisons makes it a necessary starting point for describing cognitive health among Multiracial adults, who are often excluded or recategorized in clinical and cohort studies.

Several mechanisms may explain the elevated subjective cognitive decline prevalence among Multiracial older adults. First, prior studies have linked perceived discrimination – including forms based on race, gender, or other marginalized identities – to subjective cognitive decline, though findings are inconsistent and the underlying mechanisms remain unclear.^22–24^ In addition to these well-studied forms of discrimination, Multiracial people may also experience *monoracism*, a distinct form of racism that questions, devalues, and/or delegitimizes Multiracial identities and experiences.^25,26^ In one experimental study, Franco and coauthors found that Multiracial adults randomized to recount an instance of monoracism had prolonged elevation in systolic blood pressure compared to controls,^27^ suggesting that monoracism may act as a chronic stressor with potential downstream cognitive effects.

Second, even in the absence of explicit experiences of discrimination, Multiracial individuals may struggle with healthy racial identity development, which may create unique cognitive and emotional burdens and potentially long-term cognitive vulnerability. Multiracial individuals often navigate shifting racial perceptions, identity ambiguity, and difficulty finding belonging in monoracial communities, which may require constant identity negotiation, self-monitoring, and code-switching.^28–30^ While research directly linking identity-related cognitive effort to dementia risk is limited, emerging evidence suggests that chronic stress and psychological vigilance can contribute to allostatic load, mental health vulnerability, and reduced cognitive reserve across the life course.^31–33^ Indeed, the elevated prevalence of mental health disorders, substance use, social isolation, and loneliness observed among Multiracial adults and adolescents may reflect long-term consequences of early-life and ongoing effortful coping with monoracism and identity-related social stressors.

Finally, the high prevalence of subjective cognitive decline among Multiracial adults may partially reflect group composition effects. National data show that the largest Multiracial subgroup is those identifying as American Indian or Alaska Native,^34^ a group that our study and others have found exhibits elevated subjective cognitive decline prevalence.^17^ Running Bear and colleagues’ analysis of BRFSS data^35^ found that American Indian or Alaska Native adults identifying as Multiracial had similar or worse health across multiple chronic disease, functional limitation, and health care access indicators compared to those identifying as monoracial. These findings suggest that Multiracial American Indian or Alaska Native individuals may experience intersectional structural disadvantage related to both Multiracial and Indigenous identity, which could contribute to elevated subjective cognitive decline burden in this subgroup. Shared social, structural, and intergenerational exposures could therefore contribute to similar cognitive health disparities in these overlapping populations, highlighting the importance of disaggregation and intentional racial categorization decisions in aging and dementia research.

National surveillance systems like BRFSS are critical for identifying early signals of cognitive health disparities in emerging population groups. Although we pooled five years of BRFSS data to increase statistical power, only 1.5% of our study sample (adults aged 50 and older) identified as Multiracial, compared to 10% of U.S. adults in the 2020 Census. This discrepancy likely reflects cohort effects; the youngest participants in our sample were born in 1973, just six years after the *Loving v. Virginia* Supreme Court decision that struck down the final state-level bans on interracial marriage. Because the social, legal, and institutional conditions that enabled the growth and recognition of the Multiracial population have shifted only recently, the proportion of older adults identifying as Multiracial remains relatively small but is expected to grow substantially in the coming years.^36^

Despite these strengths, surveillance systems like BRFSS still have limitations. For example, our analysis is constrained by the race and ethnicity coding in the publicly available national BRFSS, which precludes further disaggregation of the Multiracial category to investigate heterogeneity of risk across Multiracial subgroups (e.g., Asian-Black, American Indian or Alaska Native-White, etc.). One potential strategy is to request BRFSS data directly from health departments in states that release further-disaggregated race and ethnicity data,^37^ although this approach would further limit analytic sample sizes. Alternatively, surveillance systems such as BRFSS could facilitate such analyses at the national level by further standardizing the coding of race and ethnicity in state-level data collection processes to enable subgroup disaggregation in publicly available data.^38^ As the Multiracial older adult population grows, it will be increasingly important to ensure their inclusion in longitudinal and clinical dementia studies that incorporate objective cognitive assessments, which can complement and strengthen findings based on self-reported subjective cognitive decline.

Progress toward that goal has been limited by longstanding gaps in race and ethnicity measurement and by a lack of participant diversity in datasets with detailed cognitive, diagnostic, and biomarker data related to Alzheimer’s disease and related dementias. National surveys (e.g., BRFSS, the National Health Interview Survey) include Multiracial participants, but investigators have typically excluded them from analyses or reassigned them into residual categories.^15–17^ Some administrative and clinical datasets (e.g., Medicare claims data, the Chicago Health and Aging Project, Atherosclerosis Risk in Communities Study) do not include Multiracial participants or lack a comparable race category, while researchers using datasets where some Multiracial participants may be present (e.g., in the Health and Retirement Study) have also not reported estimates for this group.^4–7^ These practices, combined with inconsistent racial classification schemes, prevent deeper examination of cognitive health among older Multiracial adults. While sample sizes may remain small in individual studies, harmonizing race and ethnicity data collection across cohorts and aligning with federal standards could enable pooled analyses to determine whether the elevated subjective cognitive decline burden observed in BRFSS corresponds with differences in clinical cognitive performance, biomarker profiles, or progression to Alzheimer’s disease and related dementias. Even limited inclusion of Multiracial participants in well-characterized cohorts and clinical datasets would support more nuanced investigations of risk and resilience patterns in this growing but understudied population.

Given established links between subjective cognitive decline and dementia risk—and the likelihood that self-reported measures such as subjective cognitive decline may underestimate the true population burden of cognitive impairment^13,18^ – our results underscore the need to investigate structural and social factors shaping cognitive health disparities in the Multiracial population. These group-specific factors remain poorly characterized in both population surveillance and cohort studies. Funding agencies and research institutions should prioritize efforts to recruit Multiracial participants into dementia and life course studies, and ensure that data collection practices and research designs evolve to meaningfully include this fast-growing and diverse population. Greater inclusion of Multiracial and other historically understudied groups in cognitive aging research is essential to improving equity in dementia prevention and cognitive health promotion for all.

## Data Availability

Data used for this analysis are available online at https://www.cdc.gov/brfss/annual_data/annual_data.htm

---

**Table.** Participant characteristics and subjective cognitive decline status^a^ by self-identified race/ethnicity, BRFSS 2019-2023

|  | **Overall**<br>N = 546371<br>(100%) | **AIAN**<br>N = 7085<br>(1.1%) | **Asian**<br>N = 9012<br>(3.5%) | **Black**<br>N = 40979<br>(11%) | **Hispanic**<br>N = 28906<br>(12%) | **Multiracial**<br>N = 9347<br>(1.5%) | **NHPI**<br>N = 1487<br>(0.2%) | **White**<br>N = 431372<br>(67%) | **Other race**<br>N = 3592<br>(0.7%) | **Unknown**<br>N = 14591<br>(2.9%) |
| --- | --- | --- | --- | --- | --- | --- | --- | --- | --- | --- |
| **Sex** |  |  |  |  |  |  |  |  |  |  |
| Female | 304,670<br>(53.2%) | 3,833<br>(48.8%) | 4,656<br>(48.8%) | 25,162<br>(54.7%) | 15,974<br>(51.9%) | 5,064<br>(56.0%) | 815<br>(52.2%) | 240,387<br>(53.7%) | 1,727<br>(51.6%) | 7,052<br>(47.4%) |
| Male | 241,701<br>(46.8%) | 3,252<br>(51.2%) | 4,356<br>(51.2%) | 15,817<br>(45.3%) | 12,932<br>(48.1%) | 4,283<br>(44.0%) | 672<br>(47.8%) | 190,985<br>(46.3%) | 1,865<br>(48.4%) | 7,539<br>(52.6%) |
| **Age Category** |  |  |  |  |  |  |  |  |  |  |
| 50-54 | 65,894<br>(17.0%) | 1,182<br>(19.9%) | 1,693<br>(21.9%) | 5,999<br>(21.0%) | 6,457<br>(23.6%) | 1,552<br>(19.8%) | 283<br>(24.7%) | 46,577<br>(14.9%) | 444<br>(14.3%) | 1,707<br>(15.7%) |
| 55-59 | 74,653<br>(16.0%) | 1,283<br>(20.2%) | 1,421<br>(21.7%) | 6,525<br>(17.9%) | 5,600<br>(19.9%) | 1,511<br>(16.5%) | 276<br>(18.6%) | 55,674<br>(14.7%) | 480<br>(15.5%) | 1,883<br>(15.4%) |
| 60-64 | 88,528<br>(18.4%) | 1,395<br>(19.9%) | 1,401<br>(18.5%) | 7,414<br>(19.7%) | 5,124<br>(18.7%) | 1,689<br>(19.4%) | 296<br>(21.3%) | 68,403<br>(18.1%) | 584<br>(19.8%) | 2,222<br>(16.5%) |
| 65-69 | 92,639<br>(14.8%) | 1,192<br>(15.2%) | 1,429<br>(14.6%) | 7,076<br>(13.9%) | 4,327<br>(13.3%) | 1,493<br>(14.7%) | 225<br>(8.8%) | 73,857<br>(15.2%) | 570<br>(13.9%) | 2,470<br>(15.6%) |
| 70-74 | 86,500<br>(13.5%) | 894<br>(9.8%) | 1,268<br>(11.6%) | 6,046<br>(12.4%) | 3,323<br>(10.7%) | 1,318<br>(11.6%) | 184<br>(12.2%) | 70,660<br>(14.4%) | 529<br>(13.2%) | 2,278<br>(12.5%) |
| 75-79 | 64,021<br>(9.5%) | 565<br>(6.9%) | 866<br>(6.9%) | 3,840<br>(7.4%) | 2,161<br>(7.0%) | 902<br>(10.1%) | 117<br>(7.5%) | 53,427<br>(10.4%) | 395<br>(10.3%) | 1,748<br>(10.5%) |
| 80+ | 74,136<br>(10.8%) | 574<br>(8.1%) | 934<br>(4.8%) | 4,079<br>(7.8%) | 1,914<br>(6.6%) | 882<br>(7.9%) | 106<br>(6.8%) | 62,774<br>(12.3%) | 590<br>(13.0%) | 2,283<br>(13.8%) |
| **Subjective Cognitive Decline** |  |  |  |  |  |  |  |  |  |  |
| Yes | 69,827<br>(13.2%) | 1,210<br>(18.7%) | 939<br>(9.5%) | 5,570<br>(13.5%) | 3,929<br>(14.0%) | 1,542<br>(19.2%) | 200<br>(12.9%) | 53,948<br>(12.9%) | 574<br>(18.2%) | 1,915<br>(13.1%) |
| No | 476,544<br>(86.8%) | 5,875<br>(81.3%) | 8,073<br>(90.5%) | 35,409<br>(86.5%) | 24,977<br>(86.0%) | 7,805<br>(80.8%) | 1,287<br>(87.1%) | 377,424<br>(87.1%) | 3,018<br>(81.8%) | 12,676<br>(86.9%) |

## Notes

### Competing Interest Statement

The authors have declared no competing interest.

### Clinical Protocols

https://github.com/lamhine/brfss_scd

### Funding Statement

No funding was received for this report. TLH reports other funding from the Training Program in the Stanford Division of Endocrinology, Gerontology and Metabolism T32 DK007217-48 and subaward funding from NIH-NCI U54 CA280811. BDJ reports other funding from NIH-NIA R01AG072559. Other funders were not involved in any aspects of this study's conceptualization, design, analysis, or interpretation of results.

### Author Declarations

IRB of Stanford University waived ethical approval for this work.

