## Supplement for "Prevalence of subjective cognitive decline among older Multiracial adults, 2019-2023"

**Methods Supplement**

Additional methodological details to support the STROBE checklist are described below:

- **Study design**: This study employed a cross-sectional design to estimate crude and adjusted period prevalence (2019-2023) of subjective cognitive decline (SCD) nationally, stratified by race and ethnicity.
- **Setting**: The Behavioral Risk Factor Surveillance System (BRFSS) is a cross-sectional survey of adults ages 18 and over administered annually by US state and territorial health departments with support from the Centers for Disease Control and Prevention (CDC). BRFSS collects information on health-related determinants, behaviors, chronic conditions, and access to care through interviews on landline telephones and cell phones. Jurisdictions use probability-based sampling for participant selection. Participant recruitment into BRFSS is on-going, and there is no longitudinal participant follow-up. Additional information about the BRFSS design is available at: <https://www.cdc.gov/brfss/annual_data/2023/pdf/Overview_2023-508.pdf>
- **Participants**: Eligible participants are non-institutionalized adults (18 and over) residing in the US and participating US territories.
- **Informed consent**: Informed consent was obtained verbally from all participants included in the BRFSS.
- **Variables**: The variables used in the main analysis were (1) subjective cognitive decline, (2) 5-year age categories, (3) sex, and (4) self-identified race/ethnicity categorized by the investigator.
- **Data sources/measurement**:
  - Subjective cognitive decline: Interviewers asked participants, “During the past 12 months, have you experienced confusion or memory loss that is happening more often or getting worse?” We coded participants responding “yes” to the question about experiencing worsening or more frequent confusion or memory loss as having SCD. Participants responding “no” to the question were categorized as not having SCD.
  - Age: BRFSS reports participant age in 5-year categories (i.e., 18-24, 25-29, 30-34, etc.). We restricted our analyses to those in the following 5-year age categories: 50-54, 55-59, 60-64, 65-69, 70-74, 75-89, 80+.
  - Sex: Interviewers asked participants, “Are you male or female?”
  - Race and Ethnicity: Interviewers asked participants, “Are you Hispanic, Latino/a, or Spanish origin?” For participants indicating yes, interviewers asked “Are you Mexican, Mexican American, Chicano/a; Puerto Rican; Cuban; or “Another Hispanic, Latino/a, or Spanish origin?” Participants were allowed to select multiple responses, say they didn’t know, or refuse to respond. After asking about Hispanic ethnicity, interviewers asked participants, “Which one or more of the following would you say is your race? White, Black or African American, American Indian or Alaska Native, Asian, or Pacific Islander?” For participants selecting Asian or Pacific Islander, interviewers provided Asian (Asian Indian, Chinese, Filipino, Japanese, Korean, Vietnamese, or Other Asian) and Pacific Islander (Native Hawaiian, Guamanian or Chamorro, Samoan, or Other Pacific Islander) national origin options as well. Participants were allowed to select multiple races, say they didn’t know, or refuse to respond. There is no additional information provided in BRFSS documentation about respondents choosing any of the uninformative categories. We used the _RACE and _RACE1 variables, which report a combined race and ethnicity classification in eight mutually exclusive categories.
- **Bias**: Previous research ([Schneider et al. 2012](https://pubmed.ncbi.nlm.nih.gov/20961872/), [BRFSS Summary Quality Data Report 2018](https://www.cdc.gov/brfss/annual_data/2018/pdf/2018-sdqr-508.pdf)) has shown that BRFSS response rates are lower than other population surveys, and that minoritized racial/ethnic groups are likely underrepresented in BRFSS even after accounting for non-response adjustment weights. These patterns suggest that there could be uncontrolled sample selection bias affecting the representativeness and accuracy of estimates derived from BRFSS data. Other than applying the BRFSS-calculated (landline and cellular) complex sampling weights, we did not attempt to address any other sources of bias (e.g.: information or selection bias) in the underlying datasets provided. Potential associations between identification in these groups, SCD, and nonresponse are a limitation of this analysis.
- **Study size**: After pooling the five years (2019-2023) of BRFSS data where any of the states or territories administered the SCD module, there were in total 546,371 participants.
- **Statistical methods**:
  - Survey weight adjustment: We calculated the sample sizes for each year and computed the proportion of each year’s sample relative to the total sample size. We then multiplied the original survey weights (LLCPWT) by these proportions to obtain the adjusted weights. This adjustment accounts for the division of the dataset by year-state-module combinations and ensures proper weighting across pooled year-state-module groups in the BRFSS data.
  - Covariate adjustment: We adjusted for sex and five-year age categories using predictive marginal standardization from a survey-weighted logistic regression model. This method involves fitting a survey-weighted logistic regression model and then standardizing the prevalence estimates by predicting the probability of SCD while holding sex and age constant. This adjustment ensures that differences in SCD prevalence across racial and ethnic groups are not confounded by variations in sex and age distribution. See [Muller and MacLehose 2014 in *Int J Epidemiol*](https://pmc.ncbi.nlm.nih.gov/articles/PMC4052139/) for additional details.
  - Subgroup analyses: Our paper reports estimates stratified by race and ethnicity.
  - Missing data: We handled data via multiple imputation using the mice package in R, with 20 imputations and 20 iterations. Following others’ suggestions ([Harel et al. 2017 in *Am J Epidemiol*](https://pmc.ncbi.nlm.nih.gov/articles/PMC5860387/)) to richly parameterize multiple imputation predictor models, we incorporated additional demographic and health predictors (education, mental health, depressive disorder, smoking status, any exercise, coronary heart disease, stroke, diabetes, overweight or obese, income, binge drinking, high blood pressure, high cholesterol) in the imputation model. Subsequent analyses post-imputation ran across all 20 imputations.
